# Transmission in Latent Period Causes A Large Number of Infected People in the United States

**DOI:** 10.1101/2020.05.07.20094086

**Authors:** Qinghe Liu, Junkai Zhu, Zhicheng Liu, Yuhao Zhu, Liuling Zhou, Zefei Gao, Deqiang Li, Yuanbo Tang, Xiang Zhang, Junyan Yang, Qiao Wang

**Affiliations:** School of Information Science and Engineering, Southeast University, Nanjing, China; Erasmus School of Economics, Erasmus University Rotterdam, Rotterdam, the Netherlands; School of Architecture, Southeast University, Nanjing, China; Shing-Tung Yau Center, Southeast University, Nanjing, China

**Author notes:** Correspondence to Q. Wang.

**Keywords:** COVID-19, SARS-Cov-2, Transmission dynamics, Metapopulation network, Bayesian inference.

## Abstract

The cumulative number of confirmed cases in the United States exceeded one million on 29 April 2020, becoming the country of the most serious pandemic in the world. We proposed a model to analyze the real situation and follow-up trend of the epidemic in the US.

The proposed model divides the epidemic period into two phases, and includes three different categories of transmitters: the latent population, the documented infectious population, and the undocumented infectious population. We use metapopulation network to simulate the spread of the COVID-19 in the US, and apply the Bayesian inference to estimate the key parameters of the model. We also perform component analysis and sensitivity analysis, researching the compositions of the people with COVID-19.

The results show that the basic reproduction number in the early period of propagation is 4.06. As of April 13, 2020, only 45% (95% CI: 35% - 73%) of symptom onset cases in the United States were documented. The incubation period of COVID-19 is 10.69 days (95% CI: 10.02 – 11.74). If the current level of interventions is continued, the cumulative number of confirmed cases is expected to reach more than 1.7 million in July and continue to grow.

## 1. INTRODUCTION

In December 2019, a novel coronavirus pneumonia (novel coronavirus pneumonia) (NCP) was discovered in Wuhan, Hubei Province, China. It was caused by a new coronavirus named SARS-CoV-19 [1]. Since then, the coronavirus disease (COVID-19) [2] broke out around the world. On March 11, due to its extensive spread, the World Health Organization (WHO) declared it a pandemic [2]. As of May 5th, 2020, according to Johns Hopkins University there were 3,661,970 reported cases in 186 countries and regions [3]. With the outbreak of covid-19, different countries or regions have taken different response measures, but the number of infected people has been increasing dramatically. At the same time, the epidemic has also had a huge impact on trade flows and economic development in the world.

The United States is one of the first countries to report the confirmed cases. On January 20, the CDC confirmed that the first case was found in Washington State [4]. At first in February and early March, there were not enough reliable tests in the US, because the ability for large-scale testing was a problem, so the number of confirmed cases in the US in the early stage remained at a low level [5]. In middle March, with the improvement in testing capability, the number of confirmed cases began to increase rapidly. As of May 5th, 2020, COVID-19 has infected 1,204,351 cases in all 50 states in the US [3].

Since the basic epidemiological variables are still unknown, public health authorities in many countries and regions have focused on case detection, contacting tracing and isolation measures to reduce incidence rate and mortality caused by COVID-19 [6][7][8]. In addition, it is also important to predict the development of epidemic situation. Because the population and economic situation of each state in the US are different, the epidemic situation of each state, its response measures and effects are also different. In addition, the population flow among the states will also have an impact on each state’s epidemic situation, so the accurate prediction of the development of the epidemic in the US is a challenging problem.

Based on metapopulation network and Bayesian inference, a prediction model was proposed to analyze the pandemic development in the United States. Related researches will be introduced in the next section. Then, the details of model and method are given in the section 3. Section 4 bases on the proposed model and analysis results. Our main findings, conclusions and related discussion are combed respectively in the section 5 and 6.

## 2. RELATED WORK

In this article, we used the Bayesian inference in the transmission dynamic model which was also used in the study of Ebola epidemic before. On 2016, GIANLUCA FRASSO *et al* [9] estimated unknown parameters of SEIR model in a Bayesian framework based on the combination of the reported data and prior distribution, and they mentioned that the flexible modelling made the estimation of reproductive number robust, despite the fact of misspecification of the initial epidemic states and underreporting of the infectious cases.

On February, 2020, Joseph T Wu *et al* [10], studied on the estimation of trend of COVID-19 in mainland of China. In their work, a SEIR metapopulation model was introduced to simulate epidemic dynamics in major cities of China, and Markov Chain Monte Carlo method was used to estimate the reproductive number. In our study, we utilized their work and conducted our model in a worldwide scale.

On April, 2020, Mathias Peirlinck *et al* [11], used the global network model with the local SEIR model to estimate the outbreak dynamics both in China and the United States, the prior distributions in China, including latent period, contact period and infectious period were used to generate posterior distributions in the United States. The precise timelines and reproductive number were estimated based on these mentioned distributions in areas of China and the United States.

On April, 2020, Dayton G. Thorpe *et al* [12], applied a developed model first used by the team of Imperial College London to estimate the total infections of the United States, non-pharmaceutical interventions (NPIs) were also considered in their methods, their results indicated that the infection rate is far larger than the testing positive rate and the estimated infection rate, excluding New York city, is 2.3%.

## 3. COVID-19 TRANSMISSION MODEL AND DATA PREPARATION

### 3.1. Metapopulation and Transmission Dynamics

In order to make the model better reflect the heterogeneity of each state in the United States, we use metapopulation network to describe the state of each state. Metapopulation networks are widely applied in the analysis of the spread of complex epidemics [13][14][15][16]. Each state in the United States is considered a population. Any person in a population inevitably is in one of the following six states: the susceptibles (*S*), the latent (*L*, People who have already carried the virus but have not shown obvious symptoms), the documented communicators (*I^d^*), the undocumented communicators (*I^u^*), the confirmed cases (*C*), the recovered or dead (*Out*), which can be seen in Table 1 for more details. In any population, the transition relationships between state variables is shown in Figure 1. Considering the population flow between different populations *P_i_* and *P_j_*. A flow matrix *M* is established, in which the element *m_ij_* in row *i* and column *j* represents the number of people flowing from *P_i_* to *P_j_*, for constructing the connections of metapopulation networks. For simplifying the model as much as possible within a reasonable range, the following assumptions are proposed:

1. It is assumed that the confirmed cases and documented infectious communicators will not undergo cross population movement.
2. The proportion of people in each state of population *i* moving out from population *i* is approximately the same as that in each state of population *i*.
3. Assumed that there is no significant difference in the incubation period, infection period and the time ranging from symptom onset to being confirmed between populations.
4. Considering that the number of people recovered and dead is generally far less than the number of susceptible people, people in the state *Out* will no longer be infected.
5. People once confirmed will be under quarantine and no longer infectious.

**Table 1:** List of Symbols

|  |  |  |
| --- | --- | --- |
| *Constant* | $N$ | Total population of object of study |
| *State variable* | $S$ | Number of susceptible people |
| | $L$ | Number of latent people (in incubation period) |
| | $I^d$ | Number of documented infectious people |
| | $I^u$ | Number of undocumented infectious people |
| | $C$ | Number of confirmed cases |
| | $Out$ | Number of recovered and dead people |
| *Transition variables* | $\beta$ | Rate of the number of the susceptibles and the infected |
| | $\mu_1$ | The infection ratio between undocumented and documented transmission |
| | $\mu_2$ | The infection ratio between latent and documented transmission |
| | $x$ | Proportion of confirmed cases in the infectious population |
| | $T_L$ | Duration of latent period (incubation period) |
| | $T_d$ | Time ranging from symptom onset to being confirmed |
| | $T_I$ | Duration of infection period |

**Figure 1.**
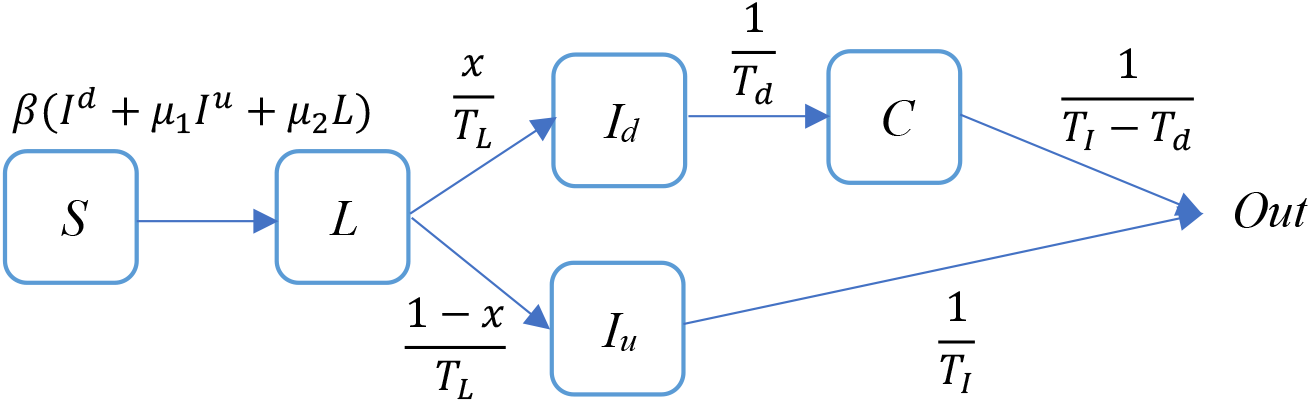
State transition diagram of transmission dynamics system.

Based on the model assumptions above, for a metapopulation model composed of *n* populations, equation 3.1 gives the transformation relationships between the variables in any population *P_i_* (*i* = 1, 2,… n).

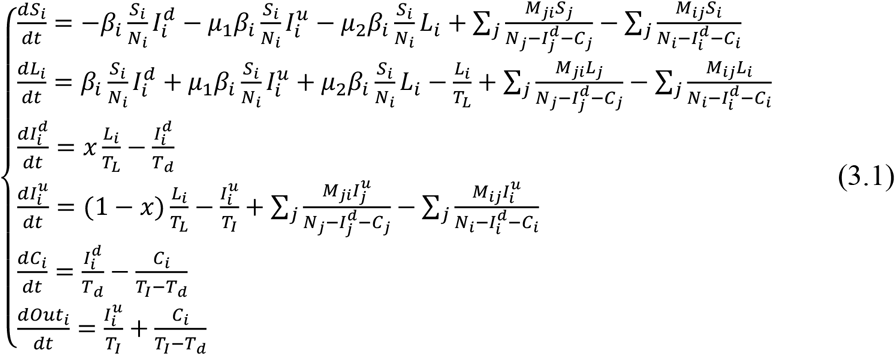

where *t* means time. *M_ij_* is the n-th order flux matrix from *P_i_* to *P_j_* (*i*, *j*=1, 2,… *n*). The other variables are explained in detail in Table 1.

### 3.2 Estimate of *T_d_* and *T_I_*

The statistical distribution test was used to estimate two important parameters: *T_d_*, the delay time ranging from symptom onset to diagnosis in patients with COVID-19, and *T_I_*, the time ranging from symptom onset to symptom disappearance (recovered or dead) in patients with COVID-19. We approximated the original distribution of *T_d_* and *T_I_* (obtained respectively from University of California, Berkeley and Johns Hopkins University respectively, see subsection 3.6 for more details) by Gamma distribution, as shown in Figure 2. The x-axis-intercept of Gamma distribution is *XI*, the estimated probability distribution of *T_d_* is Gamma (a=2.24, b=4.20, *XI*=-0.5), and that of *T_I_* is Gamma (a=2.24, b=2.50, *XI*=11). Markov Monte Carlo method is used to sample *T_d_* and *T_I_*.

**Figure 2.**
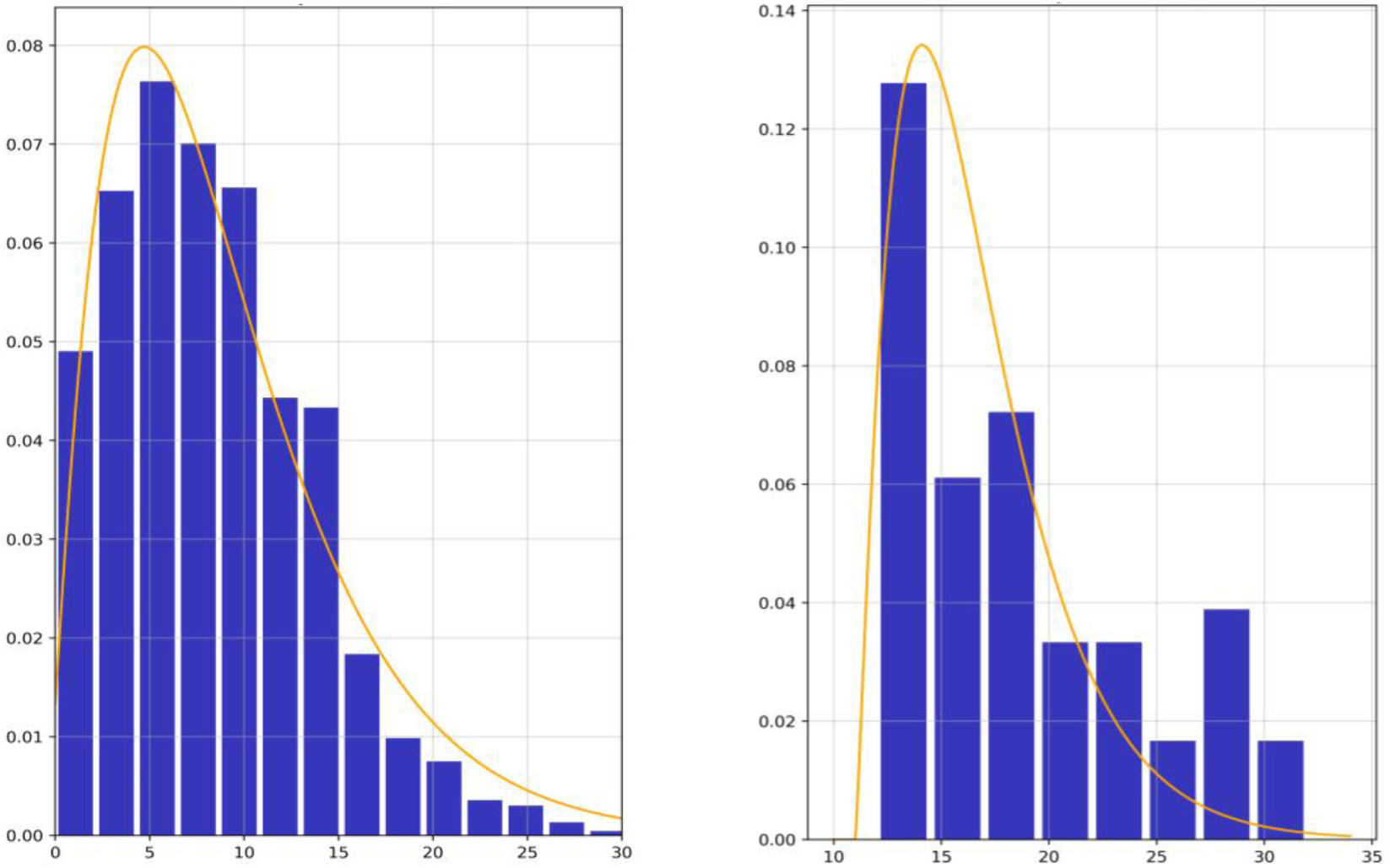
Left subgraph is the sample and probability distribution of *T_d_*, and right subgraph is the sample and probability distribution of *T_I_*. The orange curve represents the estimated probability distribution, and the blue column represents the statistical samples.

### 3.3 Practical Consideration

The COVID-19 prevention policies of major states in the United States were combed, as shown in Figure 3 (see more in appendix 1). It can be seen clearly that since March 21, 2020 (recorded as *t_0_*), many major interventions have been introduced. From this time node, the spread of the SARS-CoV-2 will be hindered by policy control and the improvement of people’s awareness of pandemic prevention. Therefore, the proposed model divides the time period into two different phases, before and after March 21, 2020. In phase one, the infection coefficient *β* is regarded as a constant, while in phase two, the *β* decreases according to the power law, as shown in equation 3.2.

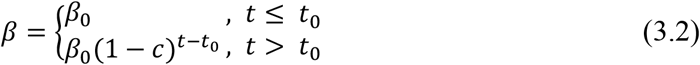

where *t* means time, *β*_0_ is a constant, and *c* is closely related to propagation blocking effect, named as the incremental inhibition ratio.

**Figure 3.**
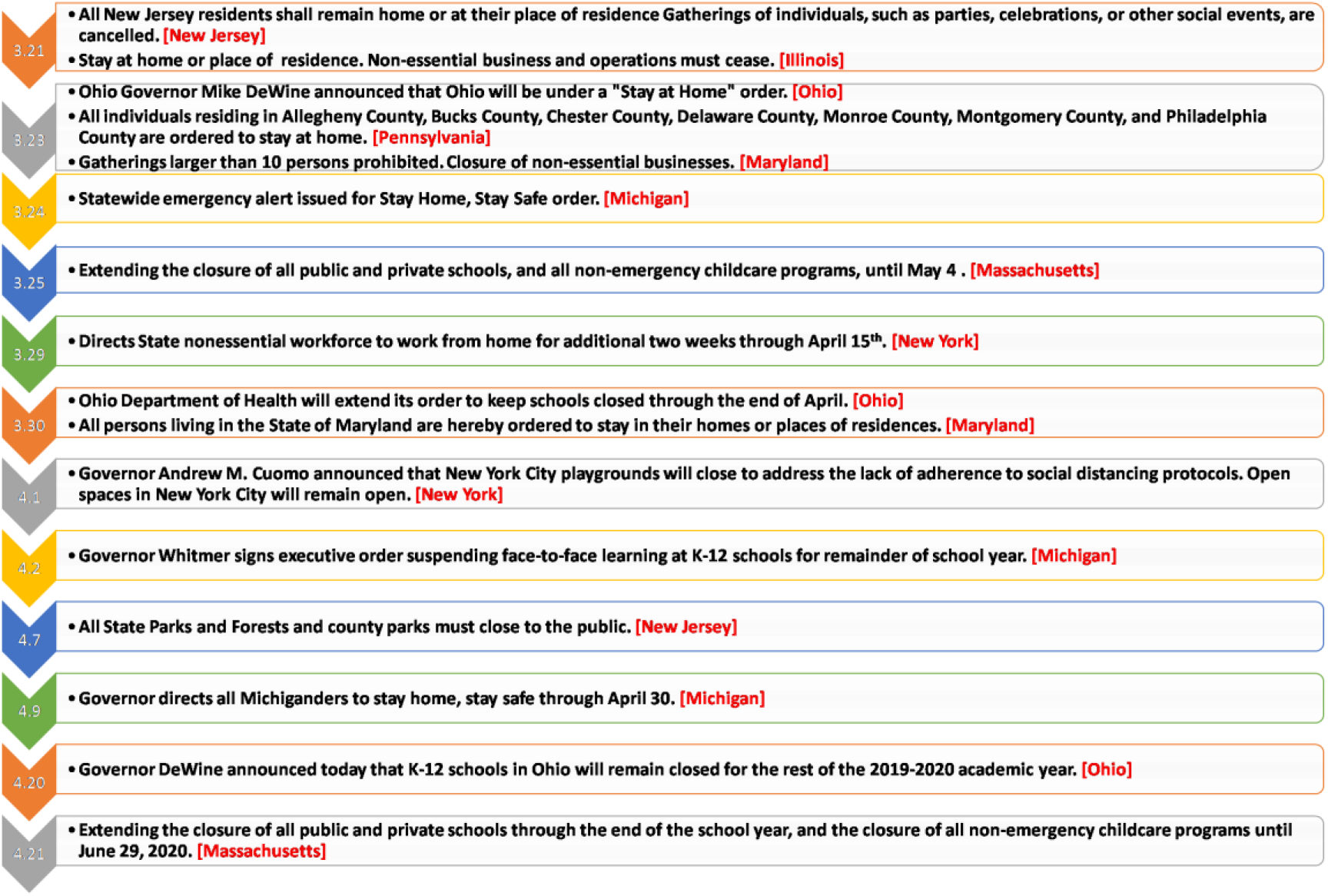
Major policies and interventions of the United States against for COVID-19.

### 3.4. Estimate of Unknown Parameters Based on Bayesian Inference

In the basic framework of Monte Carlo method, a convenient and efficient iterative solution method, Heuristic Adaptive-Step Adjustment Method (HASAM), was designed for obtaining the distribution of unknown parameters more efficiently. Its basic ideas are as follows:

Algorithm 1. HASAM

Input: The metapopulation network M, observations {*Co*} in *t* days from *start_day* to *end_day* and m states, the cumulative data after mean filtering*{cum_np_after}*, the prior distribution of ***β***, ***μ*_1_**, ***μ*_2_**, *x*, *T_L_*, the mean value of *T_I_*, *T_d_*. number of main program cycles *round_times*. The propagation dynamics of equation 3.1 is abbreviated to a function TD(·). Step adjustment factor: AF. **ε = 0.001**, used to prevent denominator from being zero.

for *round_times*=1 to *n* do

Compare the predicted value and the observed value based on the parameters extracted from the prior part, then adjust the parameters according to their gap, adjust the step size in two circles, and adjust the direction according their size relationship. The principle formula is as follows:

for *t*=*start_day* to *end_day* do

*C ←* TD(*N*, *L*, *I^d^*, *I^u^*, *C*, ***β***, **μ_1_**, **μ_2_**, *x, T_L_*)

*likelihood ← Poission*(*C, Co*)

*parameter_range* = *The range of prior distribution*

*adjust_direction* = (*Co* - *C +* ***ε***)*/|*(*Co - C +* ***ε***)*|×* (*1-likelihood*)

*adjust_length = parameter_range /* (*end_day-start_day*)*×*AF

run model with posterior obtained from the round, also compare the predicted value with the observed value, then adjust the parameter beta, the date is closer, the step size is larger. The principle formula is as follows:

*adjust_step adjust_direction*adjust_length\**(*10/*((*end_day-simu_day*) *+ 1*))

end for

the mean value of likelihood of the latest 3 days > 0.9

end for

Output: The posterior distribution of ***β***, ***μ*_1_**, ***μ*_2_**, *x*, *T_L_* while likelihood is higher than 0.9.

In every iteration of time step, the model is approaching to the highest likelihood. Compared with adjusting only once in the whole simulation time cycle, the proposed method is more efficient, but also more susceptible to noise interference. For reducing the error and making the likelihood convergence faster, we filtered the cumulative confirmed cases data. Firstly, the accumulated data was transformed into incremental data by difference, and then the 3-order mean filter was used for processing, and the time window length is 2,3,4 respectively. Finally, the incremental data was transformed into cumulative data as output.

### 3.5. Estimate of *R_0_*

*R_0_*, the basic reproduction number [17][18][19], is the average number of secondary cases that can be transmitted per unit of primary case under the condition of natural transmission. In the transmission dynamics model described in Section 3.1, the three main states that have the possibility of virus infection are: incubation period population, documented infectious population and undocumented infectious population. The durations of the three states are respectively *T_L_, T_d_, T_I_*. Their propagation ability can be expressed by intensity coefficient respectively: *μ*_2_*β, μ*_1_*β, β*. The population of a unit is always in dynamic change, if the transfer rate per unit time from the current state to the next state is 1/*T*, then the mathematical expectation of the duration can be get that 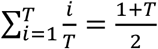. Then, *R_0_* is represented by the sum of the product of the propagatable number and the duration of each state, represented by equation 3.3.

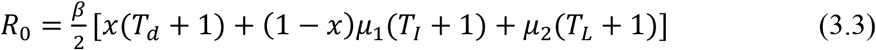

### 3.6. Data Source and Preprocessing

In this study, we used open dataset from Johns Hopkins University, Virological Organization and University of California, Berkeley. The links of the data source are shown in Appendix 1.

#### Johns Hopkins University

Johns Hopkins University shared their data on GitHub for academic and scientific research. We have compiled their open data into a chronology of confirmed data for prediction and visualization use.

#### Virological Organization

Virological Organization publishes a lot of case data accurate to individuals, including the state, location, symptoms, confirmed time, admission time and other effective information.

#### University of California, Berkeley

Researchers at the center for real estate research at the University of California, Berkeley, collated mobile signaling data flowing between states in the United States from PlaceIQ.

Because the cellular signaling data of mobile phone gives the proportion of people to be tracked moving to different places, rather than the actual number of people. Considering the equation: *number of people transferred* = *number of people tracked × transfer proportion*, and the real number of people tracked usually accounts for a certain proportion of the total local population. We used the assumed 0.001 as the scale coefficient to simulate the real number of people transferred in the algorithm design.

## 4. MAIN RESULTS AND ANALYSIS OF COVID-19 IN THE UNITED STATES

### 4.1. Parameters Estimation

Based on the equation 3.3 in section 3.5 and confirmed cases data ranging from January 21, 2020 to March 21, 2020, the basic reproduction number in the early period of propagation in the United States is estimated to be 4.06 (95% CI: 1.86 – 6.73), shown in Figure 4, which shows that the early spread of SARS-CoV-2 was very fast in the United States, and would cause a large number of cumulative cases. In order to further analyze the causes of early transmission in the United States, the normalized contributions to *R_0_* for three different categories of communicators were estimated, including L, *I^d^, I^u^*. The results showed that class *L* contributes 16.17% (95% CI: 12.86% - 21.60%) to *R_0_, I^d^* contributes 55.13% (95% CI: 43.15% - 63.97%) and *I^u^* contributes 28.70% (95% CI: 19.29% - 40.07%) to *R_0_*. It should be noted that the contribution to *R_0_* does not represent the contribution to the future pandemic spreading trend. Although the infection rate of latent period communicators is lower than that of symptomatic communicators, it still has the possibility to cause large-scale outbreak of COVID-19, which is analyzed in detail in section 4.3. Due to the fact that the documented infectious people are bound to be confirmed, most of the cases untill March 21, 2020 are infected by the confirmed cases, while 28.6% on average of the patients was infected by the undocumented infectious people, which reflects the importance to limit social distance and local contact on a large scale.

**Figure 4.**
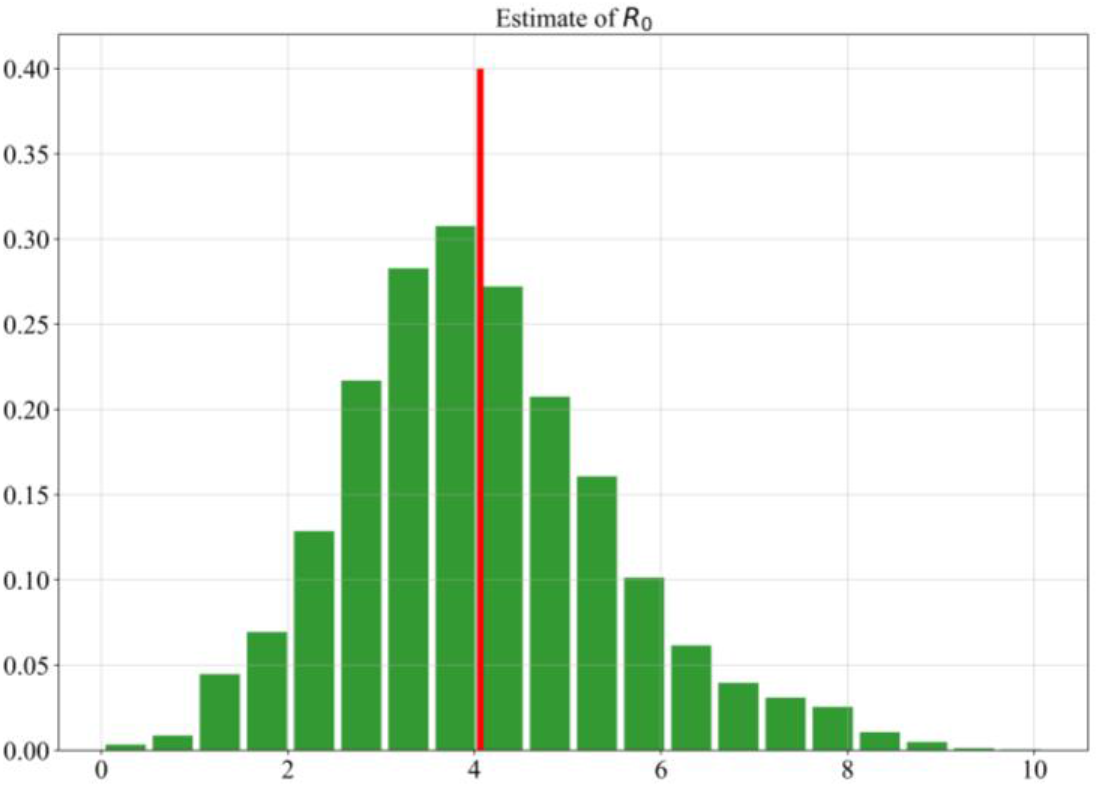
Frequency histogram of the estimated *R_0_*. The x-axis means the value of *R_0_* and y_axis means the frequency density. The red vertical line represents the mean value of *R_0_*.

The metapopulation network was used to simulate the true spread of COVID-19 in the United States, and the Bayesian inference described in Section 3.4 was used to estimate the key parameters: *β, μ*_1_, *μ*_2_, *x*, *T_L_* (explained in Table 1). As before and after March 21, 2020 (phase one and two), the transmission modes of the COVID-19 are quite different. The five states with the highest number of confirmed communicators per unit time, *β*, are New York (2.12, 95% CI: 2.00 – 2.24), Illinois (1.99, 95% CI: 1.97 – 2.10), Washington (1.97, 95% CI: 1.87 – 2.07), California (1.95, 95% CI: 1.86 – 2.00) and New Jersey (1.90, 95% CI: 1.78 – 1.98). Detailed results of *β* are shown in Appendix 3 and Figure 5. From the analysis of phase one shown in Figure 6, *μ*_1_ was estimated to be 0.40 (95% CI: 0.17 – 0.54), *μ*_2_ was estimated to be 0.06 (95% CI: 0.02 – 0.11), *x* was estimated to be 0.70 (95% CI: 0.55 – 0.78), *T_L_* was estimated to be 8.41 (95% CI: 6.64 – 9.42). For making the conclusions of the estimation compatible and explanatory with the data of the phase two (March 21, 2020 - April 13, 2020). We took this result as the prior data for the analysis of phase two. The detailed estimated results are shown in the Figure 7 and 8. It is noted that the incremental ratio, *c*, represents the degree of inhibition of SARS-CoV-2 transmission in phase two. The higher the *c* is, the better the blocking effect of virus transmission is. We found that the five states with the most serious contagion trend of COVID-19 are New York (c = 0.03, 95% CI: 0.01 – 0.04), New Jersey (0.04, 95% CI: 0.02 – 0.16), West Virginia (0.05, 95% CI: 0.02 – 0.17), Michigan (0.06, 95% CI: 0.02 – 0.21), and Missouri (0.06, 95% CI: 0.02 – 0.21). As of April 13, 2020, New York and New Jersey are the most serious pandemic areas in the United States. Their prevalence is high and the spread of COVID-19 has not been well controlled.

**Figure 5.**
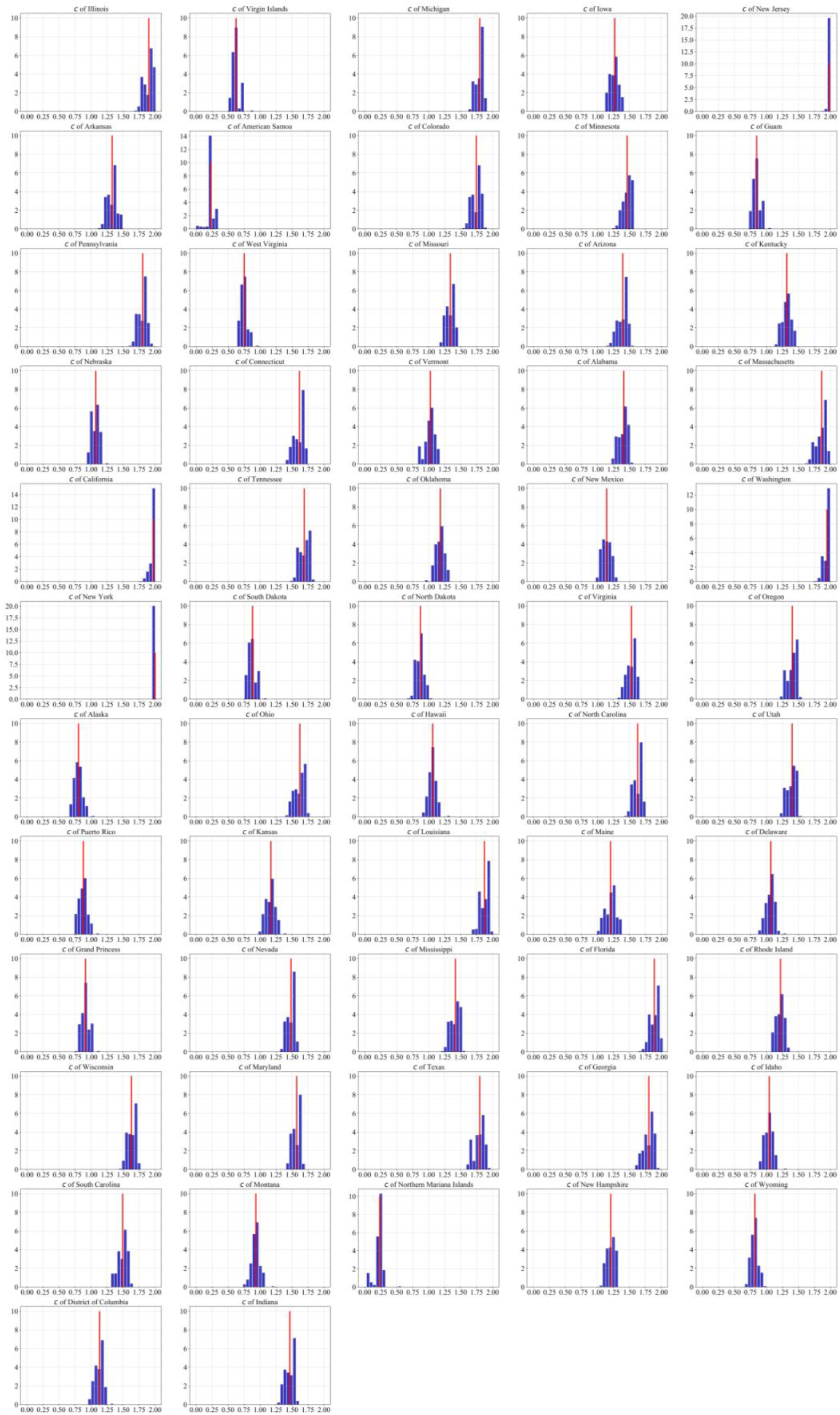
Frequency histogram of the estimated *β* in 57 states in the United States. The x-axis means the value of *β* and y-axis means the frequency density. The red vertical line represents the mean value of *β*.

**Figure 6.**
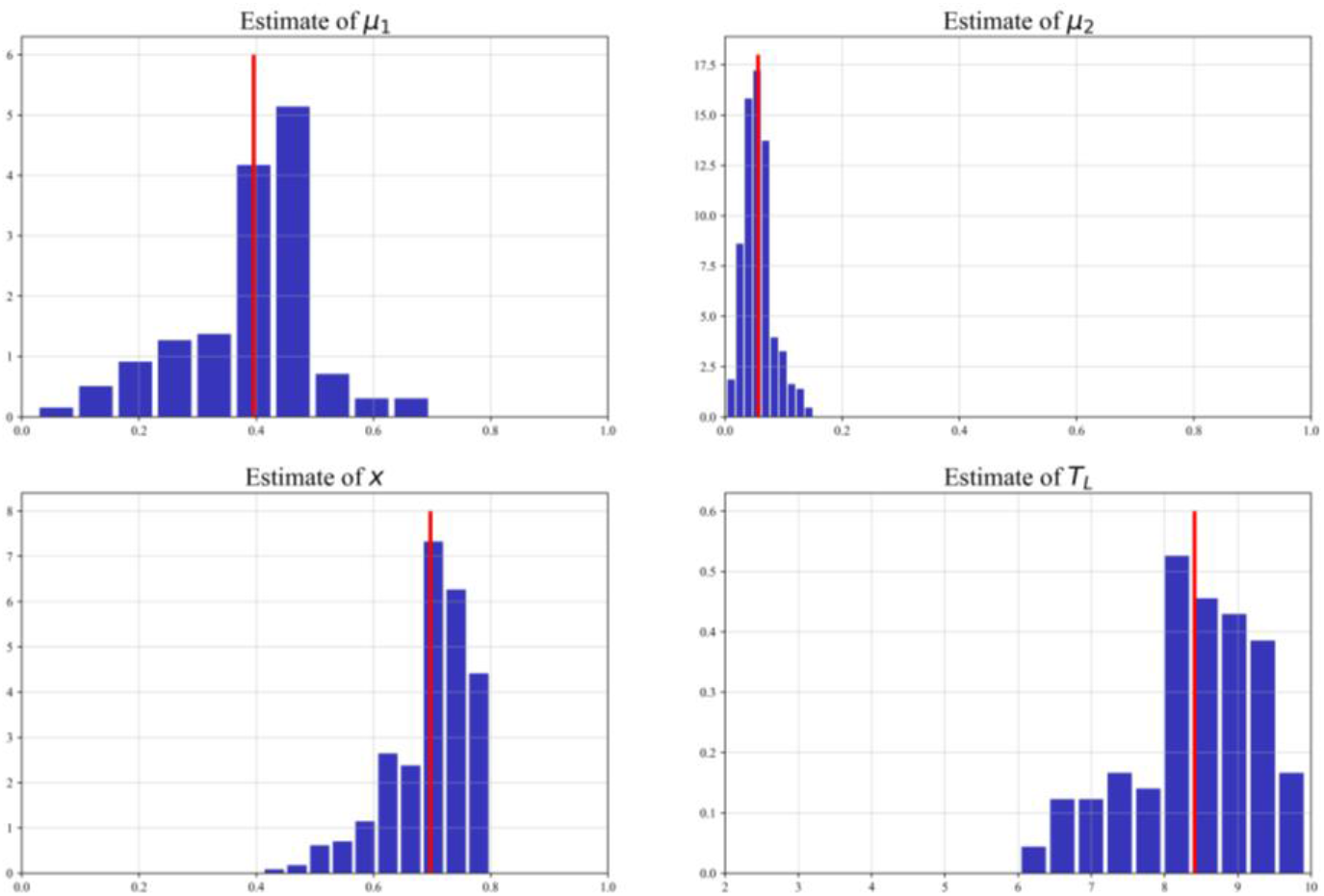
Frequency histogram of the estimated *μ*_1_, *μ*_2_, *x, T_L_* before *t_0_* The x-axis means the value and y-axis means the frequency density. The red vertical line represents the mean value.

**Figure 7.**
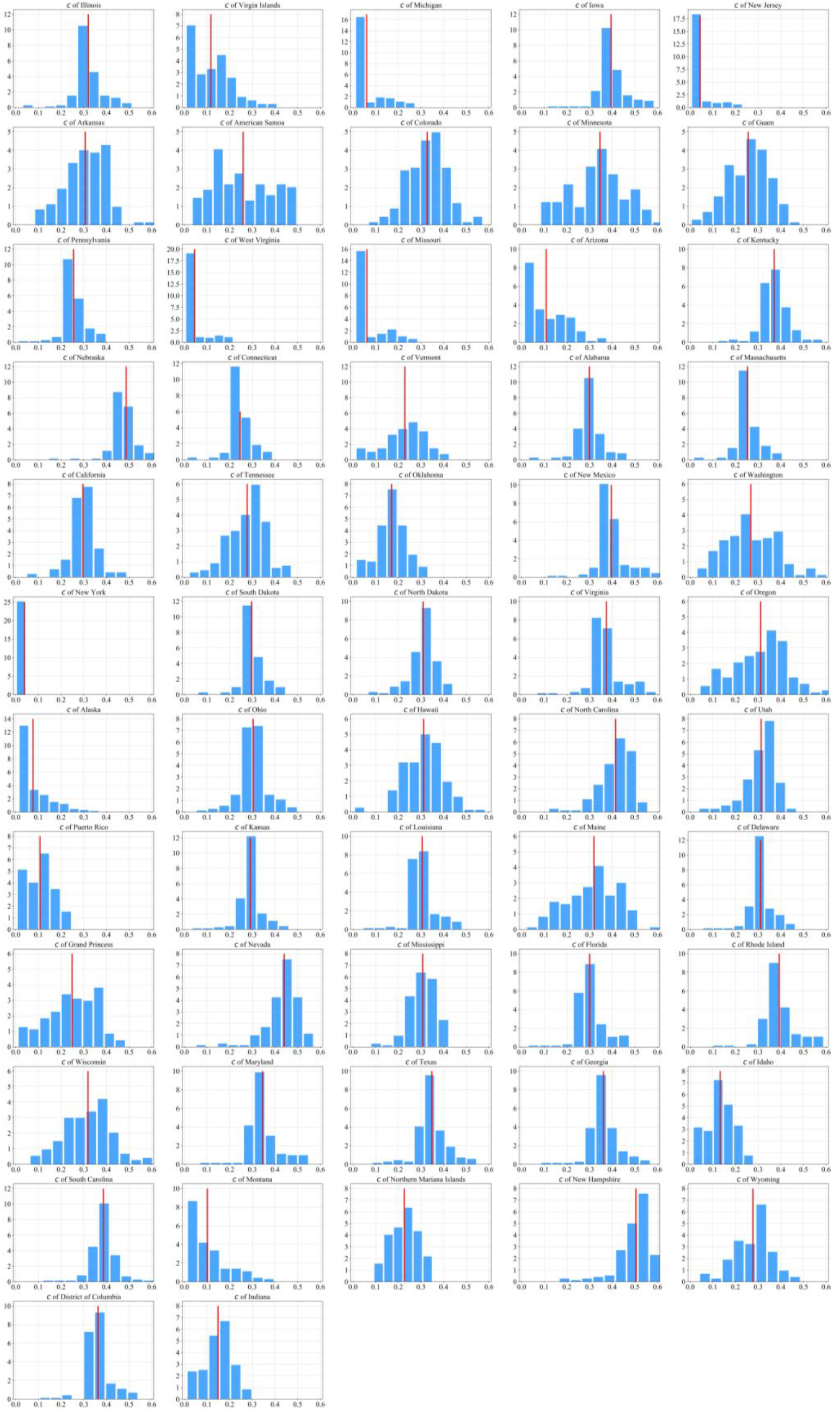
Frequency histogram of the estimated *c* in 57 states in the United States after *t_0_*. The x-axis means the value of *c* and y-axis means the frequency density. The red vertical line represents the mean value of *c*.

**Figure 8.**
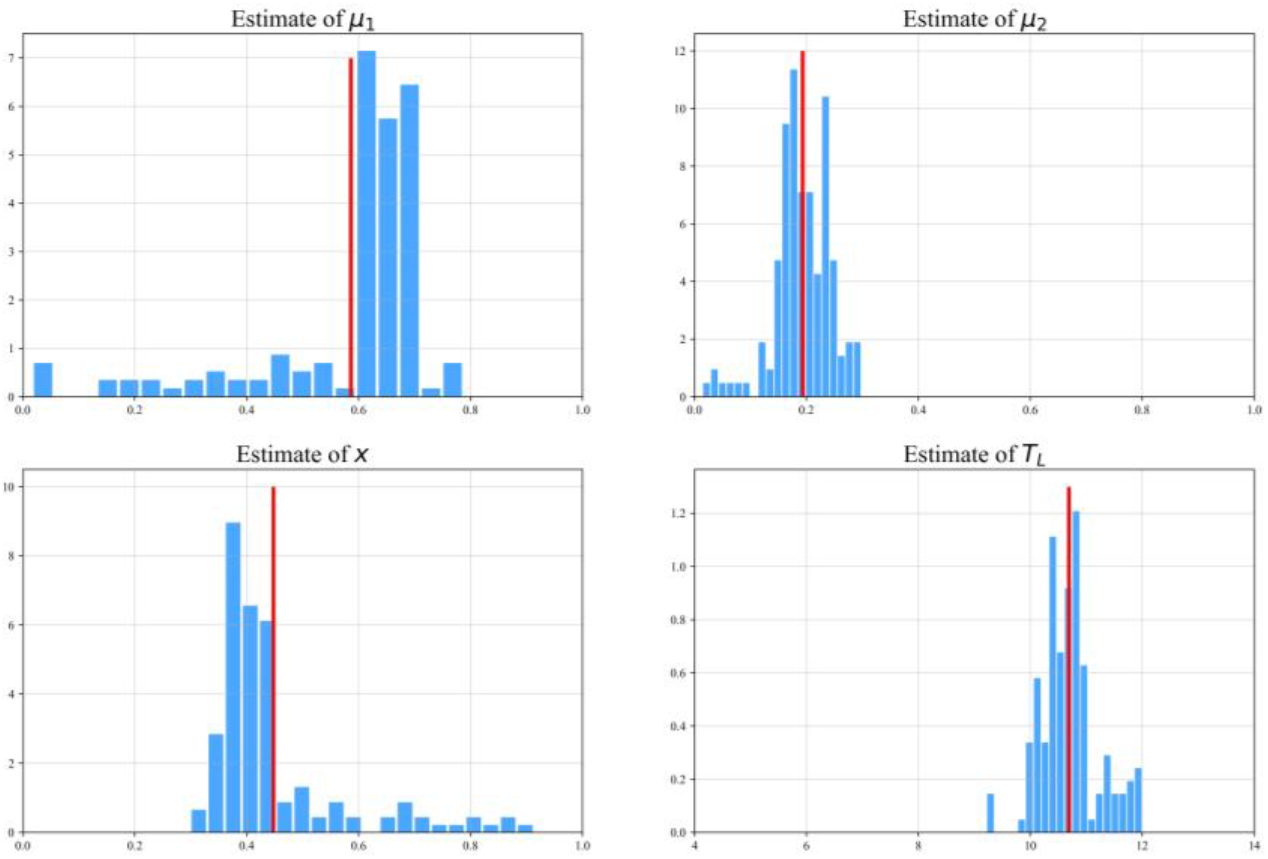
Frequency histogram of the estimated *μ*_1_, *μ*_2_, *x, T_L_* after *t_0_* The x-axis means the value and y-axis means the frequency density. The red vertical line represents the mean value.

**Figure 9.**
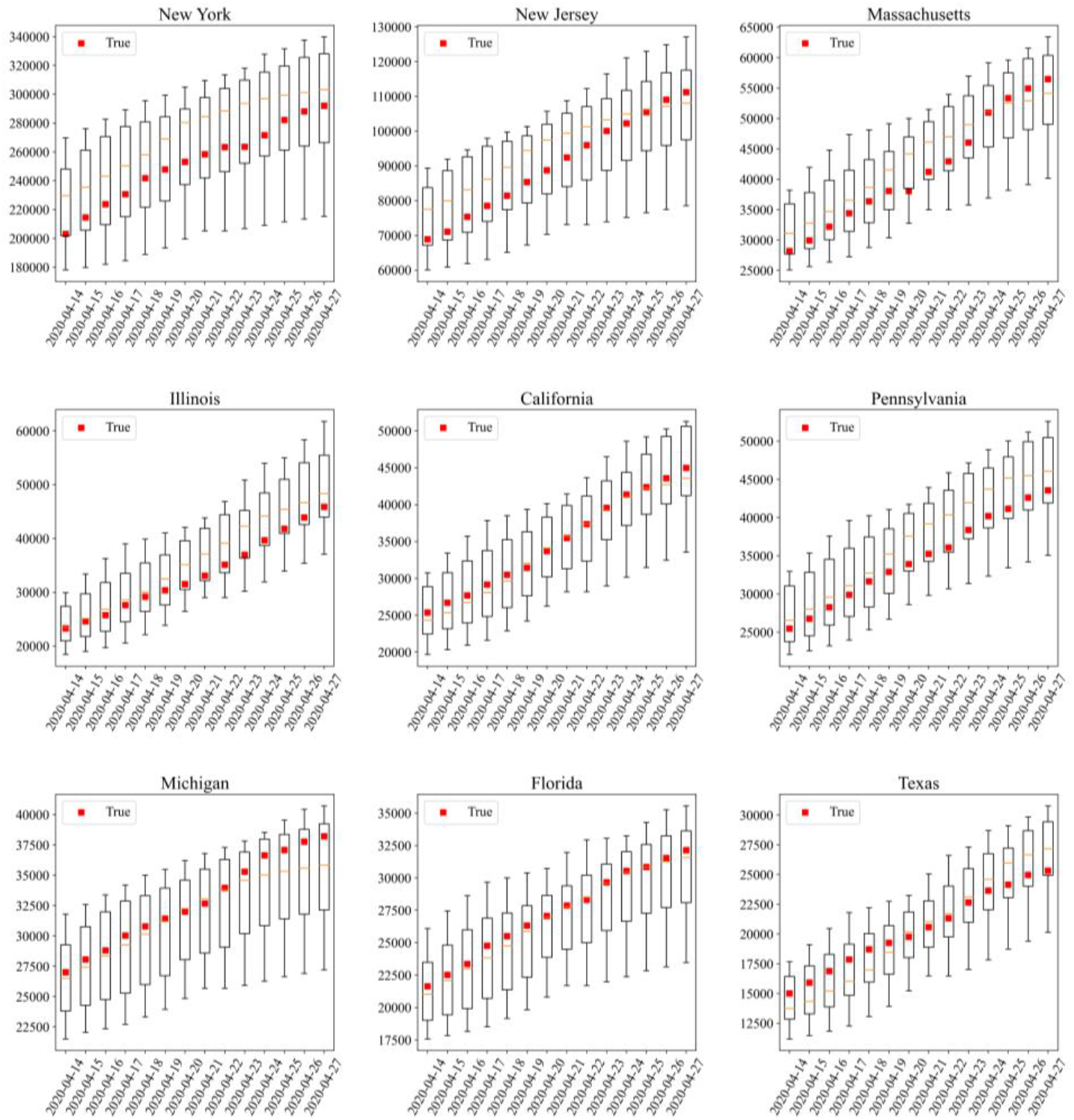
True data from several major states in the United States are compared with estimated results in the two weeks after April 13. The red blocks represent the observed true cumulative confirmed data, and the box charts represent the results of 10 independent predictions.

In fact, the epidemic situation in various regions is affected by many factors, including culture, transportation, policy, climate, etc. This shows strong heterogeneity, and it is difficult to accurately confirm a numerical result. Therefore, the prediction results only provide a more likely direction, and we pay attention to the risks and expected results in this possible direction, for warning policy makers and people to avoid them.

According to the cumulative confirmed case data from March 21 to May 3, 2020, we simulated the follow-up development of COVID-19. The results show that if the current pandemic control efforts are continued in Minnesota, Nebraska and Wisconsin, there is a possibility of serious outbreak, as shown in Figure 10. On May 3, their cumulative confirmed cases were 6663, 5661 and 7964, respectively. Although it does not show a very serious pandemic scale, the recent growth rate showed a rapid upward trend. The pandemic trend of 9 states with more severe current outbreaks in the United States were predicted in Figure 11, including New York, New Jersey, Massachusetts, Illinois, California, Pennsylvania, Michigan, Florida and Texas. Under the current situation, the cumulative number of confirmed cases in these 9 states will gradually slow down after the middle of May. If the current level of interventions are continued, the pandemic situation in the United States is likely to keep climbing up, and the cumulative number of confirmed cases is expected to reach more than 1.7 million in July and continue to grow. Considering that, the response policies of individual states in the face of epidemics are lagging behind, we adjusted the parameters of the model to force convergence of confirmed cases, as shown in Figure 12. With such modification, the number of confirmed cases in the United States eventually accumulated to about 1. 5 million. We call on all states in the United States and other regions with severe epidemics to strictly limit local contacts and reduce agglomeration. Blocking the transmission as early as possible will effectively limit the further spread of COVID-19.

**Figure 10.**
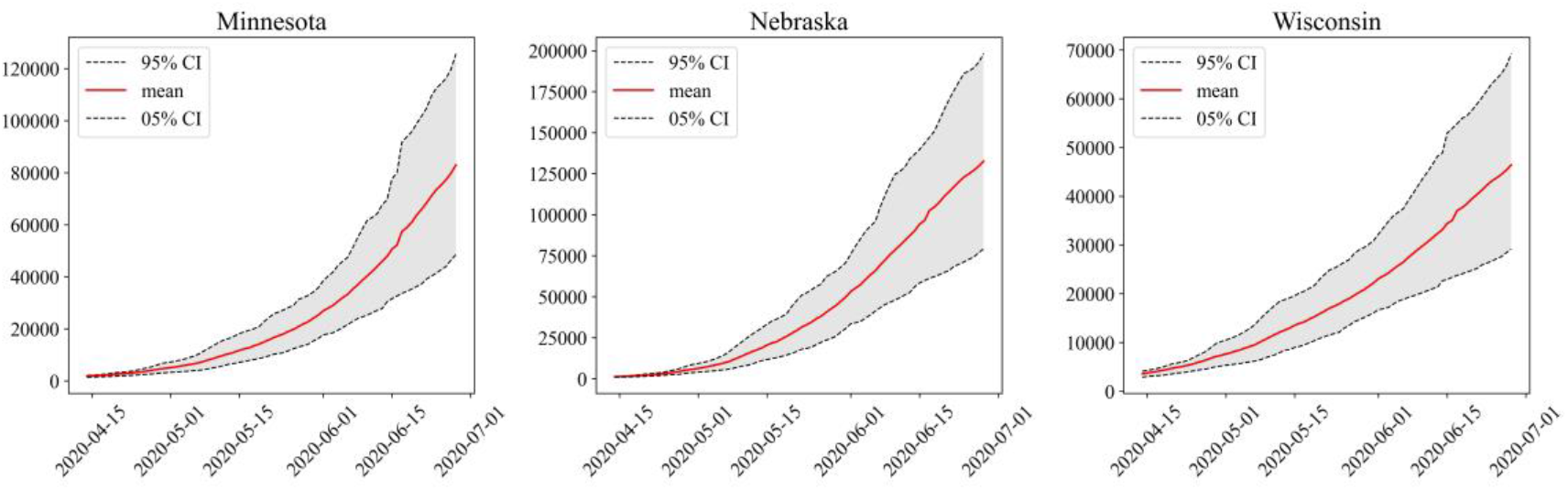
Based on the data from March 21 to April 13, 2020, the cumulative number of confirmed cases in three potentially risky states: Minnesota, Nebraska and Wisconsin, was predicted. The x-axis represents time. The y-axis represents number of people.

**Figure 11.**
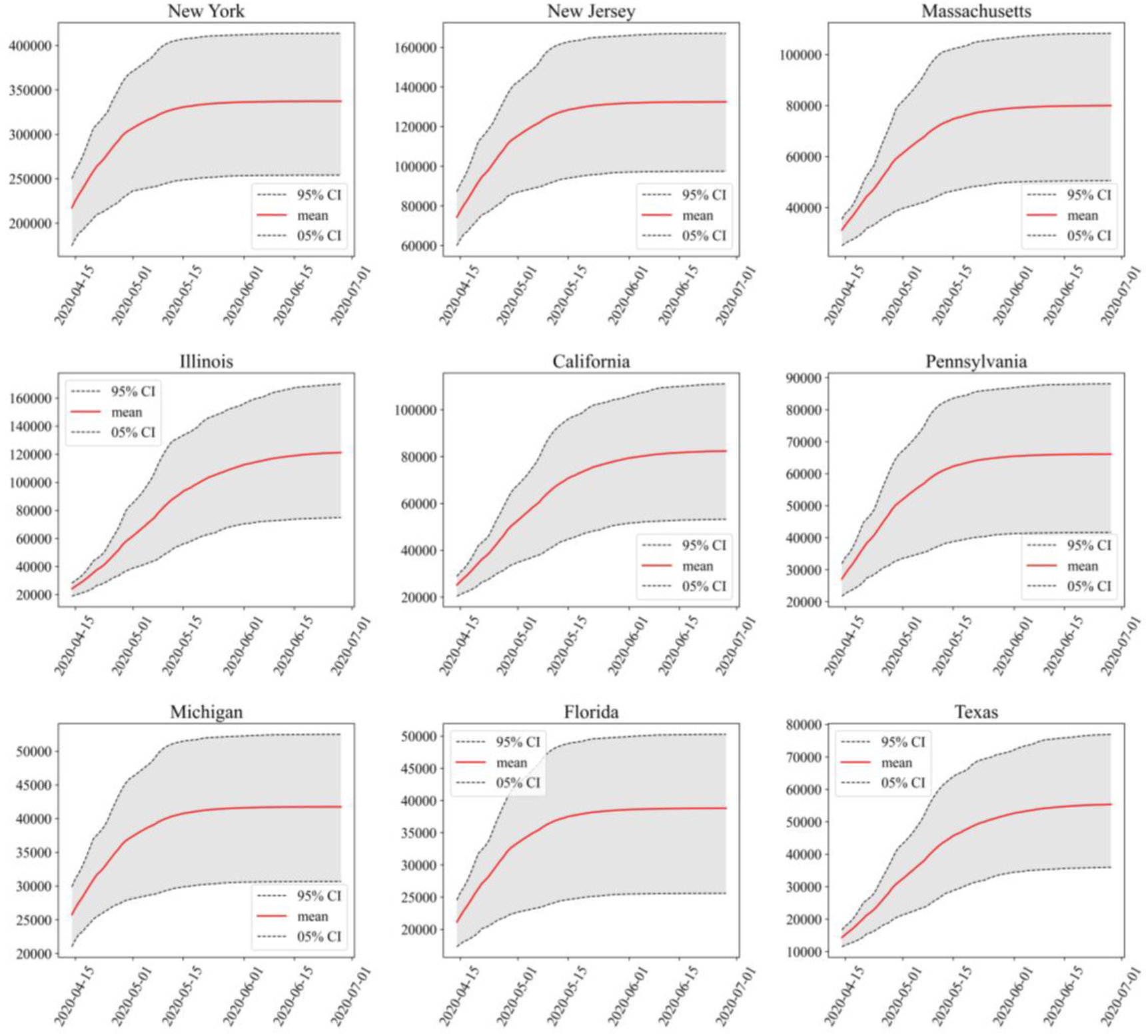
Based on the data from March 21 to April 13, 2020, the cumulative number of confirmed cases in 9 states with most severe outbreaks of COVID-19 was predicted. The x-axis represents time. The y-axis represents the number of people.

**Figure 12.**
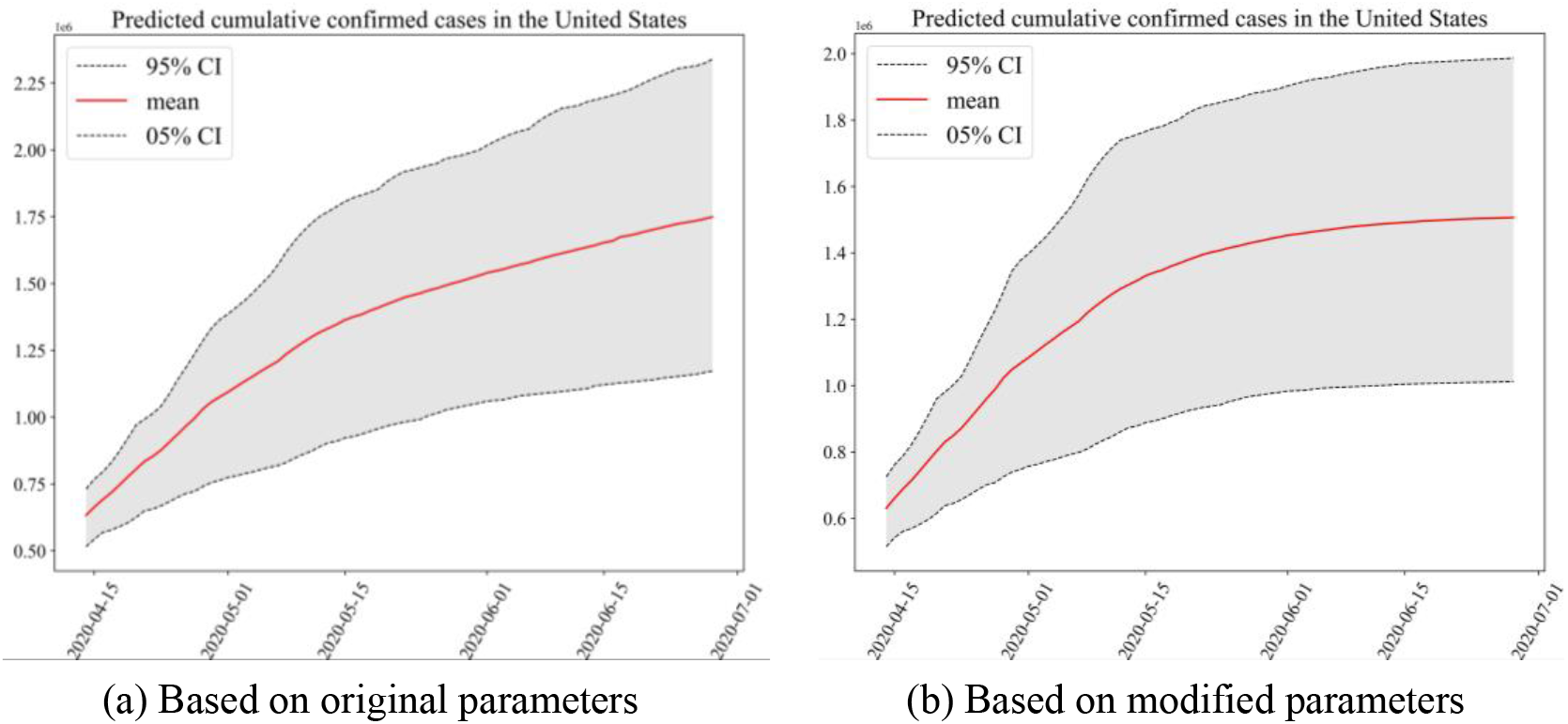
Based on the data from March 21 to April 13, 2020, the cumulative number of confirmed cases in the United States was predicted. The x-axis represents time. The y-axis represents the number of people. (a) The simulation is based on the parameters estimated by the original model. (b) The simulation is based on the estimation parameters of the model after forced suppression process. The estimated parameter *c* of Minnesota, Nebraska and Wisconsin was doubled, and *β* was reduced by 0.2.

As of April 13, 2020, it was estimated that only 45% (95% CI: 35% - 73%) of symptom onset cases in the United States have been documnented. In order to prevent more deaths due to the pandemic, the detection of COVID-19 still need to improve. The infectivity of undocumented infectious population was 0.59 (95% CI: 0.21 – 0.70) of that of the documented infectious population, while that of the latent population was 0.19 (95% CI: 0.11 – 0.27) of that of the documented infectious population. The incubation period of COVID-19 was estimated to be 10.69 days (95% CI: 10.02 – 11.74). As of April 13, 2020, the cumulative number of confirmed cases in the United States reached 580,619. The estimated number of symptomatic peploe at that time was about 1,379,693 (95% CI: 1,003,746 – 1,824,602). Under the influence of testing intensity and delay of diagnosis, the confirmation rate of symptomatic patients was about 42.08%. If considering people in the incubation period, the confirmation rate was only about 31.11%.

### 4.2. The Analysis of the COVID-19 Trend in the United States

In Section 4.1, we have preliminarily explored the epidemic situation of each state, but this does not fully explain the law of change in time. In order to further identify potential risks and give corresponding warnings, this section predicts the spread trend of COVID-19 in the United States. Before the prediction analysis, we first verified the accuracy of the model. Model was trained to several states using data from before April 13 and predicted cumulative confirmed data for the next two weeks. The comparisons between predicted results and real values are shown in Figure 9.

### 4.3. Component Analysis and Sensitivity Analysis

There exists a deviation between the observed confirmed cases and the actual number of patients. For researching the compositions of the people with COVID-19, and considering that there is only a random time delay between the number of patients in the incubation period and the actual number of patients, we simulated the change of the number of patients in the incubation period of COVID-19 in the United States over time, as shown in Figure 13. According to the conclusion of subsection 4.1, the contribution of latency population to *R_0_* is 16.17% (95% CI: 12.86% - 21.60%). However, due to the large number and imperceptibility of latency patients, it is the source of infection for 55.33% (95% CI: 55.05% - 59.95%) patients, which shows that the comprehensive isolation or quarantine measures for both symptomatic and non symptomatic groups are very effective and neccessary.

**Figure 13.**
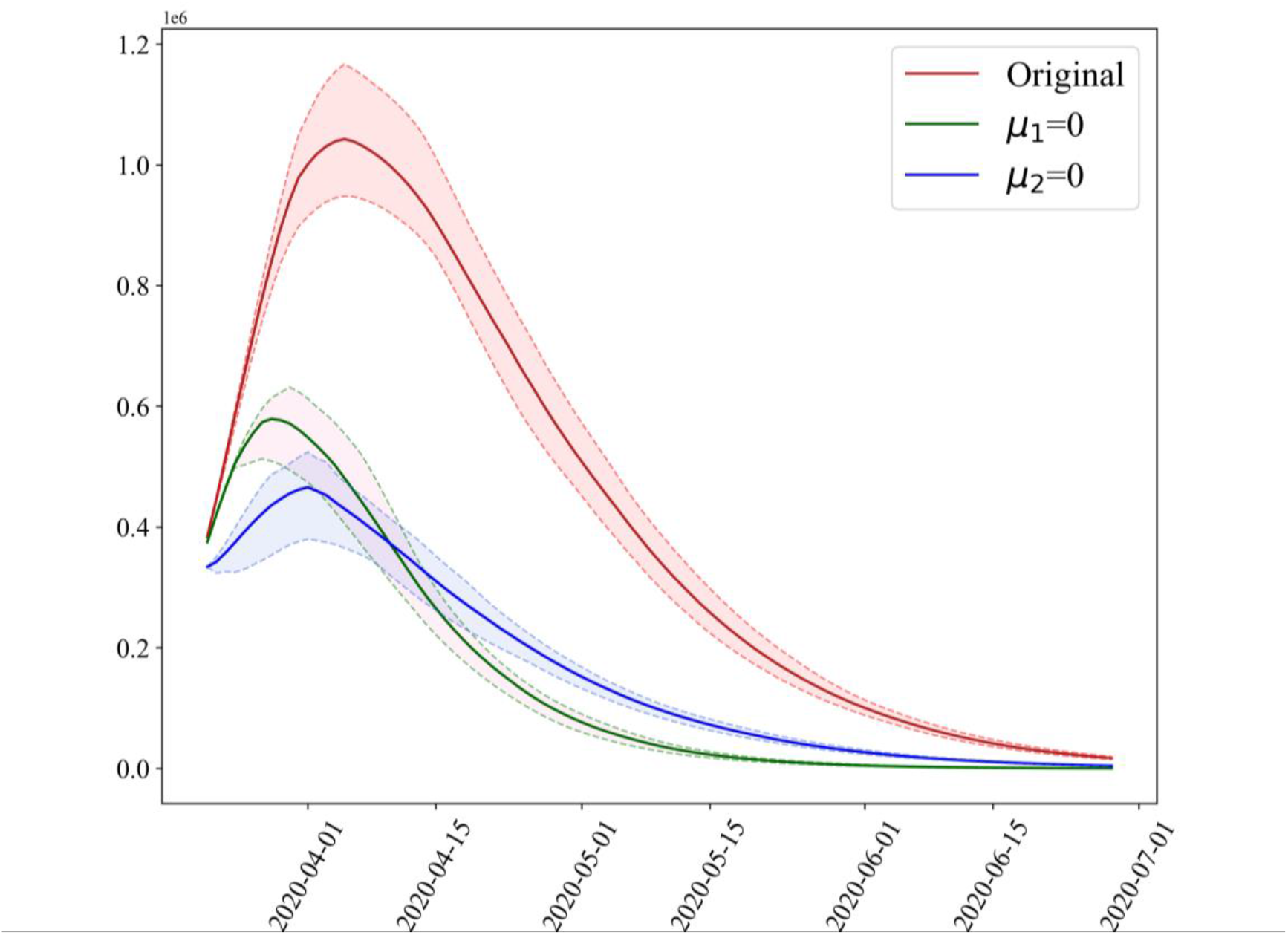
Estimates of the number of patients with incubation period over time in the United States. The x-axis represents time and the y-axis represents the number of people. The solid lines of the same color system represent the mean value, and the dotted lines represent 95% CI and 5% CI.

Effective reproduction number, *R*, is an important parameter reflecting the outbreak degree in epidemiology. Among the variables related to *R*, *β* and *T_d_* can be directly affected by human behaviours, and can change independently without greatly affecting the values of other parameters. Hence, we estimated the *R* values with *β* ranging from 0 to 0.4 and *T_d_* ranging from 0 to 60, as shown in Figure 14. The change of *R* in relation to *β* is more sensitive than that in relation to *T_d_*. The rise of *β* or *T_d_* both cause the rise of *R* value. It is worth noting that the effect of *β* on *R* is absolute influential compared with *T_d_*. When *β* = 0, propagation is terminated, *R* must be 0. While *T_d_* = 0, patients are confirmed and quarantined immediately once after symptoms onset, and the outbreak of COVID-19 is still possible (*R* > 1), so the improvement of detection intensity is effective but limited. Isolation and quarantine are more effective to pandemic prevention and control.

**Figure 14.**
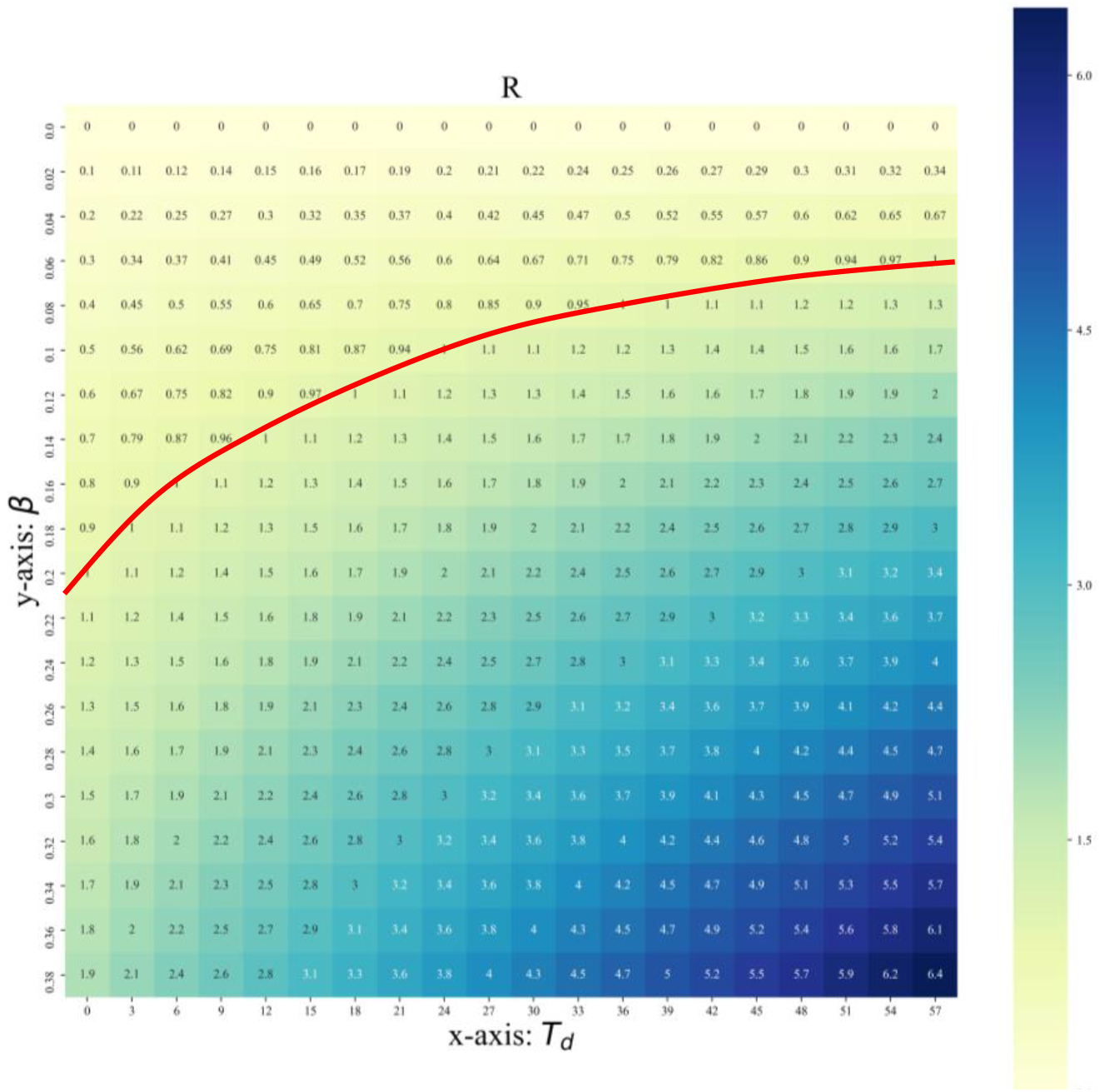
Sensitivity analysis of estimated *R* value with different *β* and *T_d_. R* at the red line position is equal to 1.

## 5. CONCLUSION

We used metapopulation network to perform a analysis of the United States based on transmission model. From the analysis, we draw the following conclusions:

1. Based on the confirmed cases data ranging from January 21, 2020 to March 21, 2020, the basic reproduction number in the early period of propagation in the United States is estimated to be 4.06 (95% CI: 1.86 – 6.73). The normalized contributions to *R_0_* for three different categories of communicators were estimated, including *L*, *I^d^, I^u^*. The results showed that class *L* contributes 16.17% (95% CI: 12.86% - 21.60%) to *R_0_*, *I^d^* contributes 55.13% (95% CI: 43.15% - 63.97%) and *I^u^* contributes 28.70% (95% CI: 19.29% - 40.07%) to *R_0_*.
2. The five states with the highest number of confirmed communicators per unit time, *β* are New York (2.12, 95% CI: 2.00 – 2.24), Illinois (1.99, 95% CI: 1.97 – 2.10), Washington (1.97, 95% CI: 1.87 – 2.07), California (1.95, 95% CI: 1.86 – 2.00) and New Jersey (1.90, 95% CI: 1.78 – 1.98). *μ*_1_ was estimated to be 0.40 (95% CI: 0.17 – 0.54), *μ*_2_ was estimated to be 0.06 (95% CI: 0.02 – 0.11), *x* was estimated to be 0.70 (95% CI: 0.55 – 0.78), *T_L_* was estimated to be 8.41 (95% CI: 6.64 – 9.42).
3. We found out that the five states with the most serious contagion trend of COVID-19 are New York (*c* = 0.03, 95% CI: 0.01 – 0.04), New Jersey (0.04, 95% CI: 0.02 – 0.16), West Virginia (0.05, 95% CI: 0.02 – 0.17), Michigan (0.06, 95% CI: 0.02 – 0.21), Missouri (0.06, 95% CI: 0.02 – 0.21). As of April 13, 2020, New York and New Jersey are the most serious pandemic areas in the United States.
4. As of April 13, 2020, it was estimated that only 45% (95% CI: 35% - 73%) of symptom onset cases in the United States have been documnented. The infectivity of undocumented infectious population was 0.59 (95% CI: 0.21 – 0.70) of that of the documented infectious population, while that of the latent population was 0.19 (95% CI: 0.11 – 0.27) of that of the documented infectious population. The incubation period of COVID-19 was estimated to be 10.69 days (95% CI: 10.02 – 11.74).
5. The results showed if the current pandemic control efforts are continued in Minnesota, Nebraska and Wisconsin, there is a possibility of serious outbreak. Under the current situation, the cumulative number of confirmed cases in these 9 states, including New York, New Jersey, Massachusetts, Illinois, California, Pennsylvania, Michigan, Florida and Texas will gradually slow down after the middle of May.
6. It was estimated that if the current level of interventions are continued, the pandemic situation in the United States is likely to keep climbing up, and the cumulative number of confirmed cases is expected to reach more than 1.7 million in July and continue to grow.

## Data Availability

All the data is available from the open website

https://github.com/CSSEGISandData/COVID-19

http://virological.org/t/epidemiological-data-from-the-ncov-2019-outbreak-early-descriptions-from-publicly-available-data/337

https://github.com/COVIDExposureIndices/COVIDExposureIndices

https://www.governor.ny.gov/news/video-audio-rush-transcript-amid-ongoing-covid-19-pandemic-governor-cuomo-announces-new-york

https://www.governor.ny.gov/news/amid-ongoing-covid-19-pandemic-governor-cuomo-announces-nys-pause-functions-extended-next-two

https://nj.gov/infobank/eo/056murphy/pdf/EO-118.pdf

## 6. DISCUSSION

In this paper, we mainly estimated and analyzed the transmission trend of COVID-19 in the United States based on metapopulation network and Bayesian inference. We estimated some parameters that were not researched in previous studies, such as the latent, undocumented, documented transmittors, etc. In fact, though some potential factors were considered in our model, the real trend in the United States is related to multiple factors and complex policies. The COVID-19 prevention policies of major states in the United States were since March 21, 2020, however, the implementation effect and efficiency of these policies are still unknown. In addition, in the two phases of epidemic transmission, except the metapopulation network, other factors, such as seasonal temperature, catering facilities, and customs or habits of each region remains to be studied.

All the above analyses are based on the assumption that there is no big deviation or error in the official statistics. In fact, there may be biases in the norms of statistical data in various states or deviation in the case data accurate to individuals, which may also lead to bias in our results.

## Abbreviations

COVID-19: Coronavirus Disease 2019; SARS-CoV-2: Severe Acute Respiratory Syndrome Coronavirus 2; CI: Confidence Interval; SEIR: Susceptible-Exposed-Infectious-Recovered.

## Declarations

### Ethics approval and consent to participate

Not applicable.

## Consent for publication

Not applicable.

## Availability of data

The datasets used and analyzed during the current study is available from open resources.

## Authors’ Contributions

Conceived and designed the experiments: Qinghe Liu, Junkai Zhu, Junyan Yang, and Qiao Wang.

Performed the mathematical modelling: Qinghe Liu, Junkai Zhu, and Qiao Wang.

Analyzed the data: Qinghe Liu, Junkai Zhu, Zhicheng Liu, and Yuhao Zhu.

Collect the data: Zefei Gao, Qinghe Liu, Liulingzhou and Junkai Zhu.

Performed the computations: Qinghe Liu, Yuanbo Tang, and Xiang Zhang.

Wrote the paper: Qinghe Liu, Junkai Zhu, Liuling Zhou, Zefei Gao, Deqiang Li, Zhicheng Liu, and Yuhao Zhu.

All authors read and approved the final manuscript.

## Acknowledgments

Not applicable.

## Appendix 1 Data Source (All data used in the paper is public.)

Real time confirmed cases data, recovered data, death data from Johns Hopkins University in the United States. https://github.com/CSSEGISandData/COVID-19

Data of symptoms onset and being confirmed time of cases from Virological Organization. http://virological.org/t/epidemiological-data-from-the-ncov-2019-outbreak-early-descriptions-from-publicly-available-data/337

Data of population flow based on mobile signaling from PlaceIQ. https://github.com/COVIDExposureIndices/COVIDExposureIndices

Policy information from New York State Government Website, New Jersey State Government Website, Commonwealth of Massachusetts Website, Michigan State Government Website, Illinois State Government Website, Ohio State Government Website, Maryland State Government Website, Pennsylvania State Government Website.

https://nj.gov/infobank/eo/056murphy/pdf/EO-118.pdf

https://nj.gov/infobank/eo/056murphy/pdf/EO-107.pdf

https://www.mass.gov/news/baker-polito-administration-announces-extension-of-school-and-non-emergency-child-care-0

https://www.mass.gov/news/baker-polito-administration-announces-extension-of-school-and-non-emergency-child-care-program

https://www.michigan.gov/coronavirus/0,9753,7-406-98158-525173--,00.html

https://www.michigan.gov/coronavirus/0,9753,7-406-98158-524028--,00.html

https://www.michigan.gov/coronavirus/0,9753,7-406-98158-522731--,00.html

https://coronavirus.illinois.gov/s/resources-for-executive-orders

https://ohio.gov/wps/portal/gov/site/media-center/news-and-events/covid-19-update+schools-remain-closed

https://ohio.gov/wps/portal/gov/site/media-center/news-and-events/dewine-extends-school-closure-order

https://ohio.gov/wps/portal/gov/site/media-center/news-and-events/ohio-issues-stay-at-home-order-and-new-restrictions-placed-on-day-cares-for-children

https://govemor.marvland.gov/wp-content/uploads/2020/03/Gatherings-FQURTH-AMENDED-3.30.20.pdf

https://www.governor.pa.gov/wp-content/uploads/2020/03/03.23.20-TWW-COVID-19-Stay-at-Home-Order.pdf

## Appendix 2 Simulation Results of *β* and *c* in 57 States of the United States

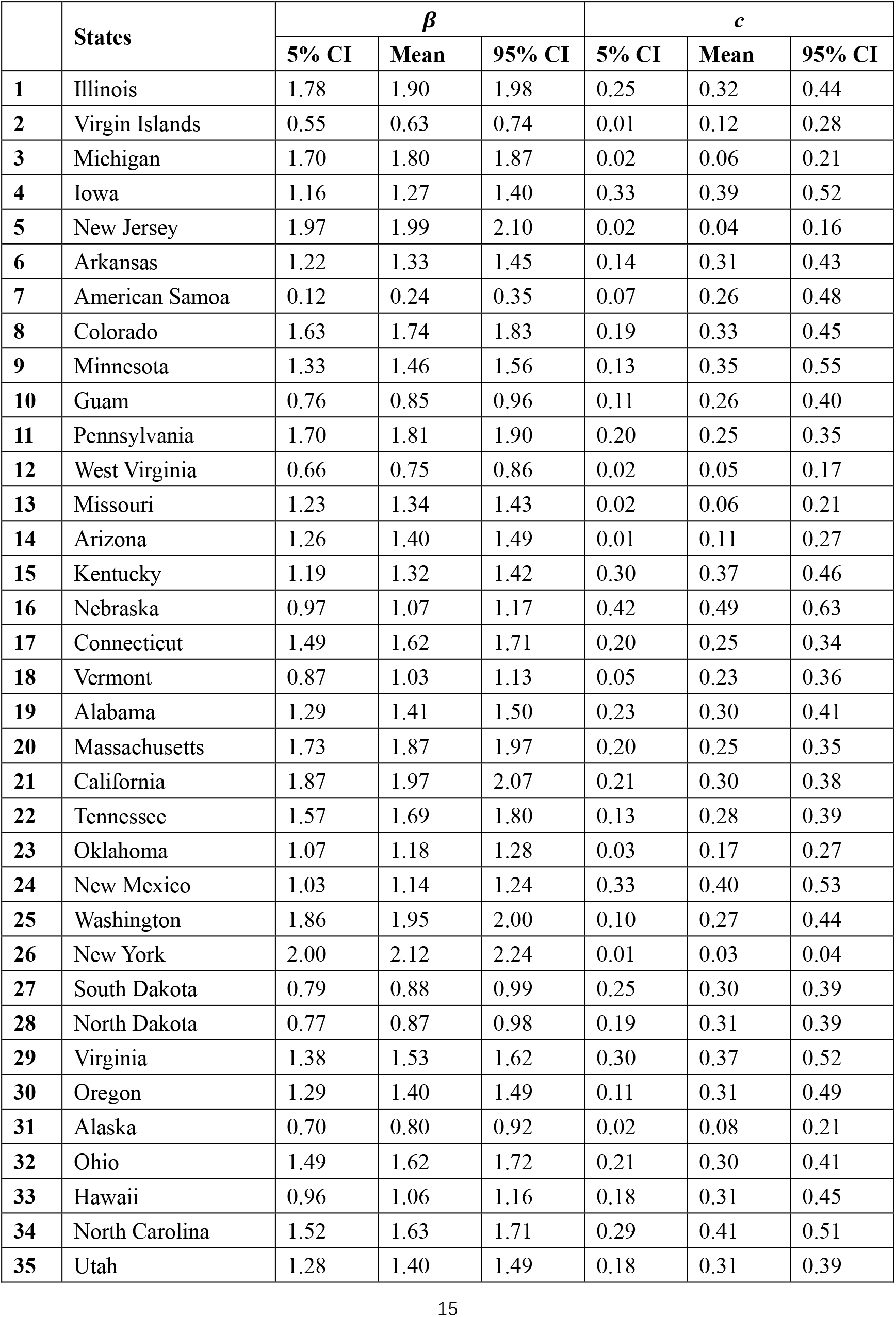
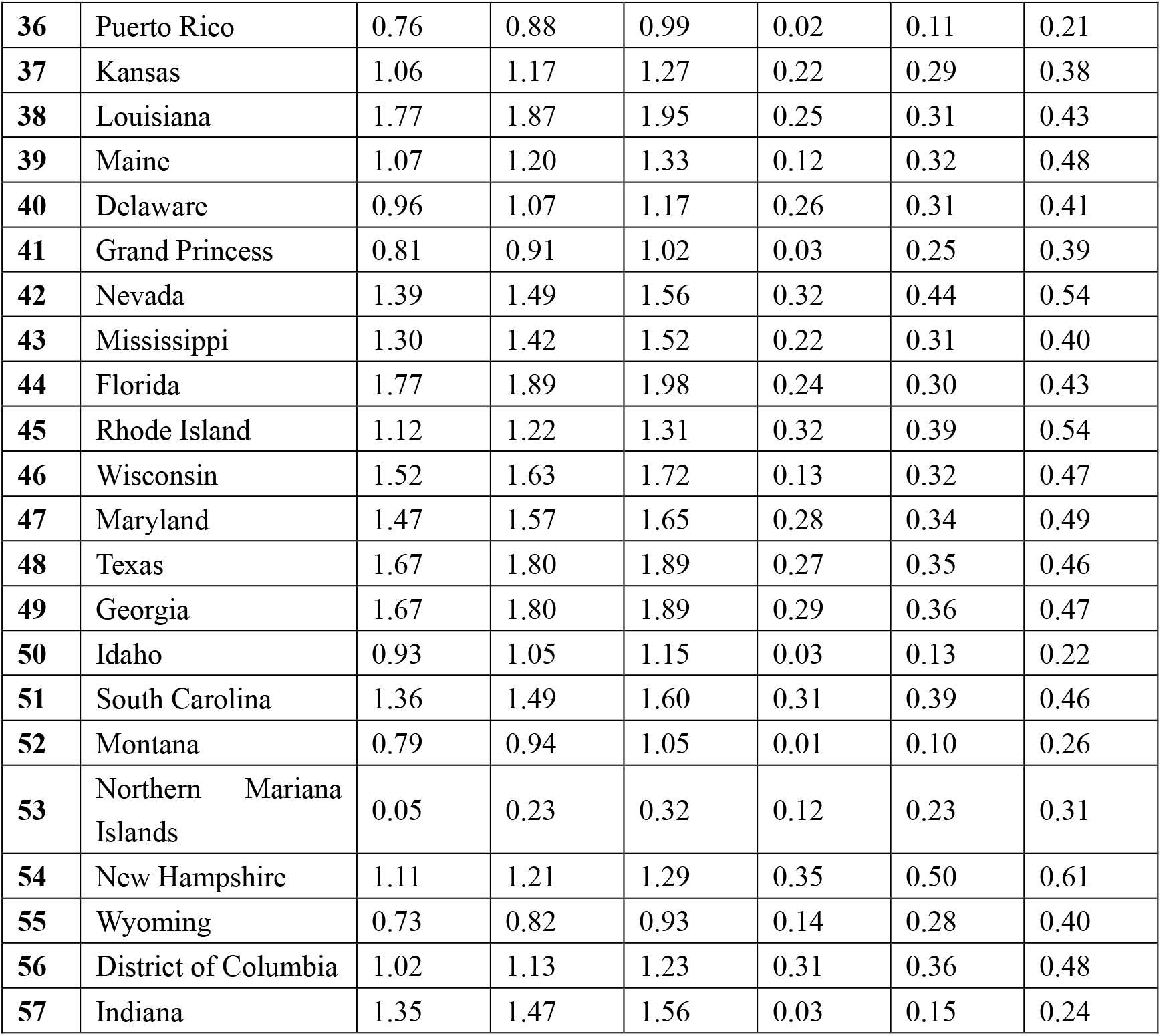

